# Resources to Aid Ethical Review of Clinical Studies: An Exploratory Scoping Review Identifying Gaps and Opportunities

**DOI:** 10.1101/2023.12.13.23299842

**Authors:** Merle-Marie Pittelkow, Daniel Strech

## Abstract

**Background:** Research Ethics Committees (RECs) review the ethical, legal, and methodological standard of clinical research. However, complying with all requirements and professional expectations while maintaining the necessary scientific and ethical standards can be challenging for applicants and members of the REC alike. There is a need for accessible guidelines and resources to help medical researchers and REC members navigate the legal and ethical requirements and the process of their review.

**Methods:** We employed an explorative search for resources on the websites of a purposively selected sample of relevant stakeholders including 12 national umbrella organizations (six German-language and six English-language), three English-language international umbrella organizations, and 16 national REC’s of major university hospitals (eight German- and eight English-language). We qualitatively mapped the identified resources onto the guiding principles of ethical clinical research and 35 related checkpoints. To describe the content of the resources we conducted a thematic analysis.

**Results:** We extracted a total of 233 resources, including templates (*n* = 134, 58.5%), guidelines/recommendations (*n* = 62, 26.6%), checklists (*n* = 23, 9.9%), tools (*n* = 5, 2.2%), flowcharts (*n* = 5, 2.2%), glossaries (*n* = 3, 1.3%), and one (0.4%) software program. We extracted 101 German and 132 English resources created between 2004 and 2023. The majority (*n =* 204; 87.6%) could be assigned to one checkpoint. The remaining 29 (12.5%) resources were considered unspecific (e.g., a checklist which documents to be submitted for a German drug trial). The specific resources are discussed per checkpoint.

**Conclusion:** While much support is available for some aspects such as participant information and informed consent forms, it is lacking in other areas such as study design, analysis, and biometrics. More support should be provided in these areas to ensure that research projects are methodologically sound. A more detailed analysis of the quality of available resources could help identify other areas of need.

## Background

Human clinical research plays a central role in the development and validation of therapies (including drugs, biologics, medical devices) and diagnostics (1). To ensure that clinical trials involving human participants meet the highest standards of research ethics, they must be reviewed by an institutional review board (IRB) or research ethics committees (RECs)^1^. Although there are some international differences in responsibilities (2), RECs generally evaluate clinical trials for ethical justification, including the risks and benefits to study participants, the informed consent documents, scientific validity, and methodological soundness including aspects of the study design and statistical analysis (3–6). Consequently, RECs are of vital importance for the ethical, legal, and methodological standard of clinical research.

The quality of the ethical review hinges on the quality of the documents provided by the applicants and the ability of members of RECs to evaluate these. The quality of the documents in turn depends on the applicant’s ability to navigate a complex landscape of ethical, legal, and methodological requirements. Members of RECs on the other hand must handle these aspects in a responsible and fair manner. Some aspects are governed by international law, others by federal or state law, and some refer to non-legal professional laws and guidelines such as the Declaration of Helsinki (4) or CIOMS guidelines (5). Furthermore, ethical judgements on the appropriateness of consent documents or risk-benefit ratios require expert knowledge and, unavoidably, include interpretive judgments.

Complying with all legal requirements and further professional expectations while maintaining the necessary scientific and ethical standards can be challenging for applicants and members of the REC alike. Consequently, there is a need for accessible guidelines and resources to help medical researchers and REC members navigate the legal and ethical requirements and the process of their review. Ideally, such guidance and resources should be easily accessible and readily available, enabling applicants to incorporate them into early study planning and ethics application drafting. s Assuming that the first point of contact for applicants prior to the ethics application is the websites of ethics committees and their umbrella organizations, their websites serve as the optimal platform for distributing resources to applicants before they submit their ethics applications.

Several types of resources could facilitate the application and review process, including checklists, templates, topic-specific guidelines, decision trees or online tools^2^. For example, checklists could ensure that all required information is included in the application avoiding unnecessary effort, such as the need to resubmit application. Templates could streamline the application and review process, for example ensuring that informed consent texts meet all legal requirements. REC members could also benefit from using REC-approved templates, for example by saving time and focusing only on the highlighted changes during the assessment.

Many umbrella organizations and RECs already offer online resources for applicants and REC members. For example, the German association of RECs (Arbeitskreis Medizinischer Ethik-Kommissionen, AKEK) or the World Health Organization (WHO) offer guidelines and templates online to aid the application process (7,8). However, these individual resources are spread across many websites, making it opaque what the existing resources offer. A scoping review in Pubmed and Google on “Resources to Aid Ethical Review of Clinical Studies” did not reveal a systematically developed overview of available resources. It is therefore unclear to what extent the available resources cover aspects of clinical research ethics.

The aim of this project is to explore and qualitatively describe the pool of available resources to answer the question of which types of resources are already available and which topics they cover. Our search is not intended to identify all available resources, but to give a qualitative, thematically saturated overview of what types of resources are commonly available. Therefore, we focused our search on the websites of a purposively selected sample of relevant stakeholders including umbrella organizations of RECs and RECs of major university hospitals and limited ourselves to German- and English-language resources. We have selected these groups because we believe that they are the first point of contact for applicants and REC members when faced with issues regarding research ethics in clinical trial as well as the process of writing or reviewing an ethics application.

## Methods

### Protocol and Registration

This dynamic, data-driven project, was not preregistered. Instead, we provide a project log and all relevant material on OSF (https://osf.io/e7dmt/).

### Search and Selection of Sources

We searched for online resources provided by national and international umbrella organisations for clinical research ethics as well as RECs of large university hospitals in Germany, the United States, and the United Kingdom (see Table 1). The project started in February 2023. We created a list of potential sources from personal experience and expertise. We then searched the websites of relevant umbrella organizations also applying backward searching, examining relevant stakeholders when they were mentioned on the websites of the umbrella organizations. Next, we searched the websites of RECs of major university hospitals. On each individual website, we first opened all subtabs linked to from the starting website. We then went through each subpage successively and searched for resources relating to ethics. The search was stopped in August 2023 when we reached saturation defined as encountering the same kind of resources (e.g., templates for informed consent) without being able to add untapped resources to the collection. A detailed log of the search including considerations and justifications for the inclusion and exclusion of stakeholders can be found on OSF (https://osf.io/usbt8). Due to the use of backwards searching the final selection of sources also included related umbrella organizations (e.g., Clinical Research Ethics Consultation Collaborative) and not only umbrella organizations or RECs.

**Table 1.** Overview of the informational sources.

| **Table 1.** Overview of the informational sources |  |  |  |
| --- | --- | --- | --- |
| **Language** | **Name** | **Identified via** | **Nr. Resources** |
| ***National umbrella organizations*** |  |  |  |
| German | Arbeitskreis Medizinischer Ethik-Kommissionen (AKEK) | Author team | 32 |
| German | Swiss Human Research Ethics Committee (Swissethics) | Author team | 45 |
| German | Forum Österreichischer Ethikkommissionen | Author team | 1 |
| German | Technologie- und Methodenplattform für die vernetzte medizinische Forschung e.V. | AKEK | 1 |
| German | Bundesinstitute für Arzneimittel und Medizinprodukte | Ethik-Kommission Westfalen-Lippe | 1 |
| German | Paul Ehrlich Institute | Ethik-Kommission Westfalen-Lippe | 3 |
| English | National Health Service (NHS) Health Research Authority | Author team | 10 |
| English | United Kingdom Research and Innovation (UKRI) | Author team, online search | 4 |
| English | Canadian Institutes of Health Research | Online search | 1 |
| English | SmartIRB | Online search | 1 |
| English | National Institute of Health (NIH) | Author team | 0 |
| English | Clinical Research Ethics Consultation Collaborative | Online search | 1 |
| ***International umbrella organizations*** |  |  |  |
| English | World Health Organization (WHO) | Author team | 13 |
| English | European Network of Research Ethics Committees (EUREC) | Online search | 1 |
| English | Ethics and New Emerging Research Institutions (ENERI) | Online search | 1 |
| ***National RECs*** |  |  |  |
| German | Ethik-Kommission Westfalen-Lippe | Author team | 4 |
| German | Ludwig Maximilian University of Munich (LMU) | Author team | 12 |
| German | Hannover Medical School (MHH) | Author team | 4 |
| German | Ärztekammer Hamburg | Author team | 0 |
| German | University of Cologne | Author team | 7 |
| German | Heidelberg University | Author team | 5 |
| German | Albert-Ludwigs-University Freiburg | Author team | 4 |
| German | University of Vienna | Author team | 0 |
| English | Stanford Center for Biomedical Ethics | Author team | 31 |
| English | Mayo Clinic | Author team | 0 |
| English | Johns Hopkins | Author team | 21 |
| English | University Hospital Bristol | Author team | 23 |
| English | Royal Stoke | Author team | 0 |
| English | Cambridge | Author team | 0 |
| English | Oxford | Author team | 0 |
| English | University Hospitals Birmingham | Author team | 9 |

### Information Sources

We identified 31 information sources (Table 1) including 12 national umbrella organizations, three international umbrella organizations, and 16 national REC’s.

### Eligibility Criteria

We limited ourselves to German- and English-language resources but employed liberal eligibility criteria. To be extracted, resources had to relate to the application for ethical approval for clinical studies. Clinical studies could be mono- or multi-site studies, trials testing medical products, drugs, or other medical interventions with human participants. Resources could be of several types (i.e., checklists, templates, flowcharts, or recommendations) and be addressed to the applicants (i.e., applied researchers), REC members, or both. We did not extract resources in the form of legal texts, tutorials for university-specific submission programs, or course syllabi. To avoid duplication, we did not extract resources if they were already extracted from a previous source.^3^

### Data Items

For each resource, we extracted: the stakeholder and their main website, country, year of publication, type of study if applicable, type of resource, the link to the resource, a description of the resource, and language. The extraction sheet was piloted with a selection of resources from the first stakeholder considered (i.e., AKEK) and adjusted accordingly.

### Data Charting Process and Synthesis

To qualitatively describe the resources, we mapped them onto the guiding ethical principles of an internationally established framework for clinical research, namely, social value, scientific validity, favorable risk-benefit ratio, fair participant selection, independent review, informed consent, respect for participants, and collaborative partnership (see Emanuel et al., 2008) and 35 related checkpoints presented in Raspe et al. (2012). The 35 checkpoints (Figure 1) provide a more detailed and praxis-oriented account of how the guiding principles are translated into the ethical review process.

**Figure 1.**
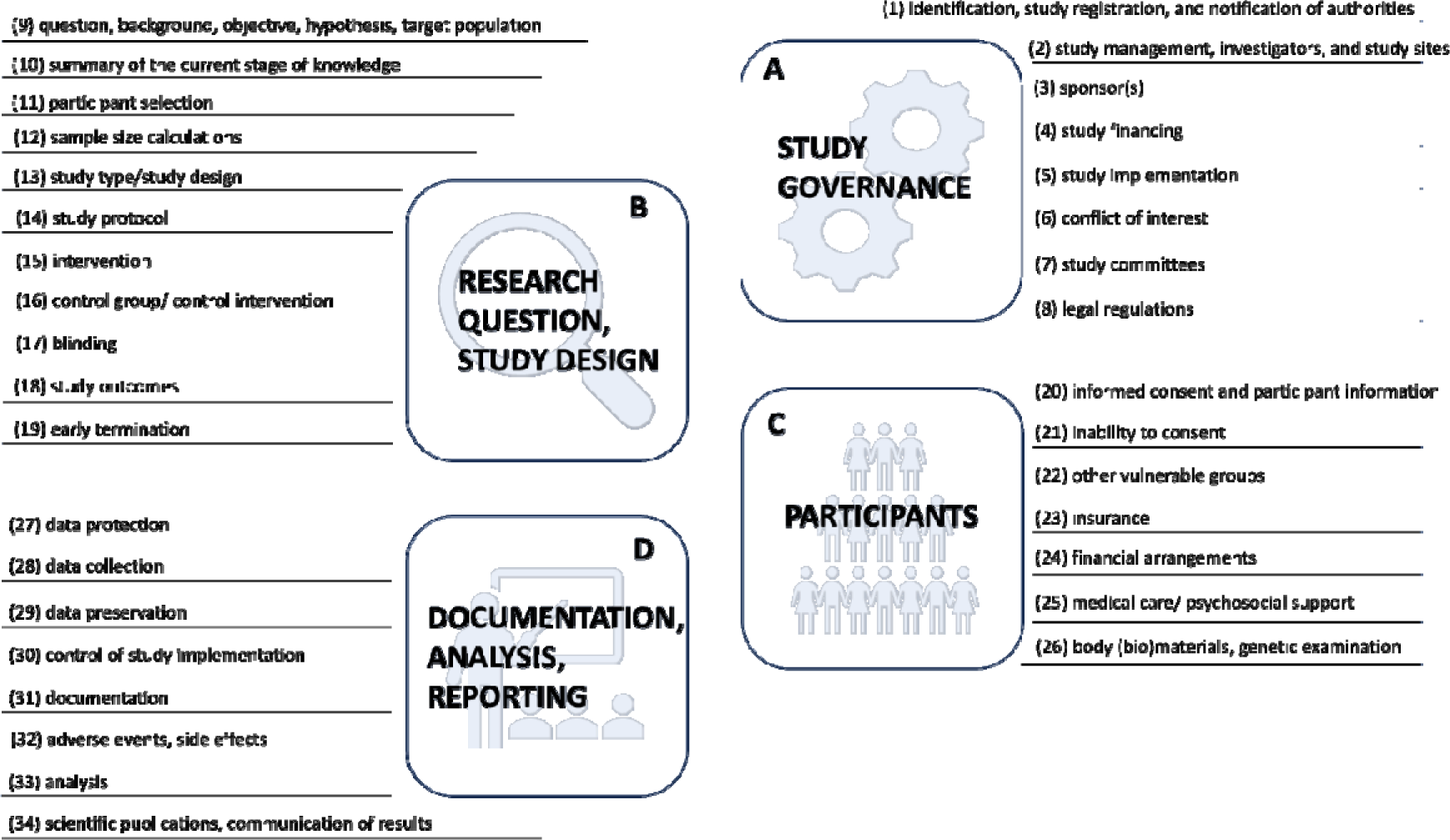
The 35 Checkpoints associated with the guiding principles of ethical clinical research.

If possible, resources were assigned to one of the 35 checkpoints during extraction. If no clear assignment was possible, the resource was grouped under ‘other’. During data analysis, we checked whether this initial assignment was congruent with the content of the resource. This resulted in 21 resources being relabeled (see the project log provided on OSF).

To describe the available resources, the first author (MMP) conducted a thematic analysis in Atlas.ti using thematic analysis as a realist method. Themes were identified at the semantic level from the content explicitly mentioned in the resources. We used a deductive approach by having the checkpoints guide our qualitative analysis and theme construction. First, documents were grouped according to the checkpoints assigned during data extraction. Next, the resources for the first ten checkpoints were coded to create a codebook. The overall structure of the codebook was continuously revised during this process. Following, the revised codebook structure (i.e., authors, target group, resource type, main topic, topic, language, study type, participant type, data type, and legal regulations) was used to code all resources. While additional subcodes were added during this stage, the overall theme structure remained.

## Results

We initially extracted 243 resources from the websites of 24 stakeholders^4^ (see Table 1) reflecting umbrella organizations of RECs and RECs of major university hospitals. During data analysis, we excluded ten resources (nine due to eligibility criteria and one duplicate). This resulted in a total of 233 resources. The majority (*n =* 204; 87.6%) could be assigned to one checkpoint. The remaining 29 (12.5%) resources were considered unspecific (e.g., a checklist which documents to be submitted for a German drug trial).

Resources included templates (*n* = 134, 58.5%), guidelines/recommendations (*n* = 62, 26.6%), checklists (*n* = 23, 9.9%), tools (*n* = 5, 2.2%), flowcharts (*n* = 5, 2.2%), glossaries (*n* = 3, 1.3%), and one (0.4%) software program. We extracted 101 German resources and 132 English resources. Resources were created between 2004 and 2023 with more resources being created or updated recently (see Figure 2.)

**Figure 2.**
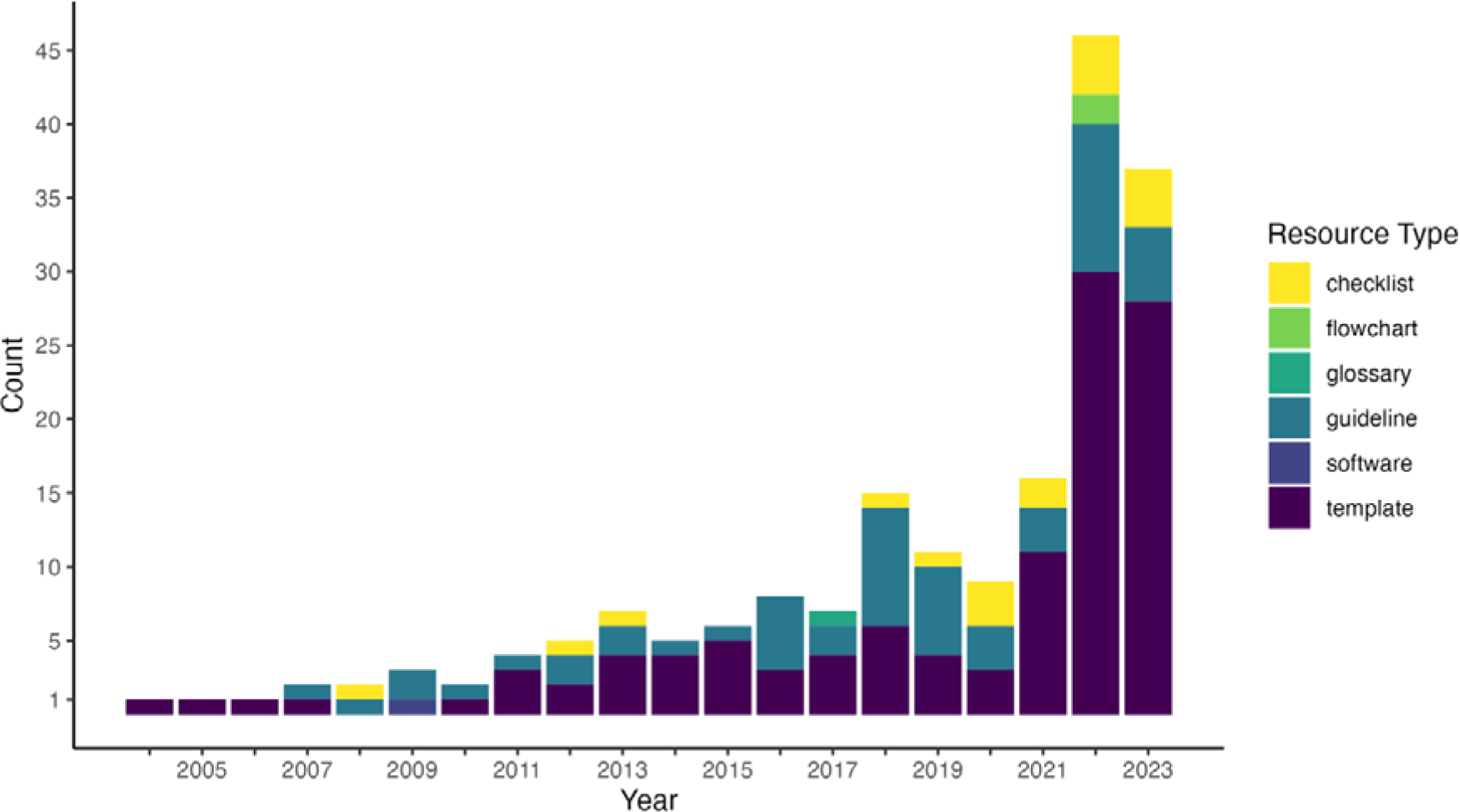
Development of resources over time. Note that for 45 resources we were unable to extract a year of creation.

### Qualitative Description of Resources

Below, we provide a brief overview of the available resources per checkpoint. A more detailed overview of the available resources per checkpoint and the themes they cover is presented in Table 2 and the full list of all available resources can be found in the supplement. Following the structure proposed by Raspe and colleagues (10) we grouped checkpoints and resources under the headings of (1) study governance/management [*n =* 24, 10.30%], (2) research question and study design [*n* = 32, 13.73%], (3) study participants [*n* = 99, 40.77%], and (4) documentation, analysis, and dissemination [*n* = 49, 21.03%] (see Figure 1). The remaining 29 (12.45%) resources were grouped under ‘other’.

**Table 2.**
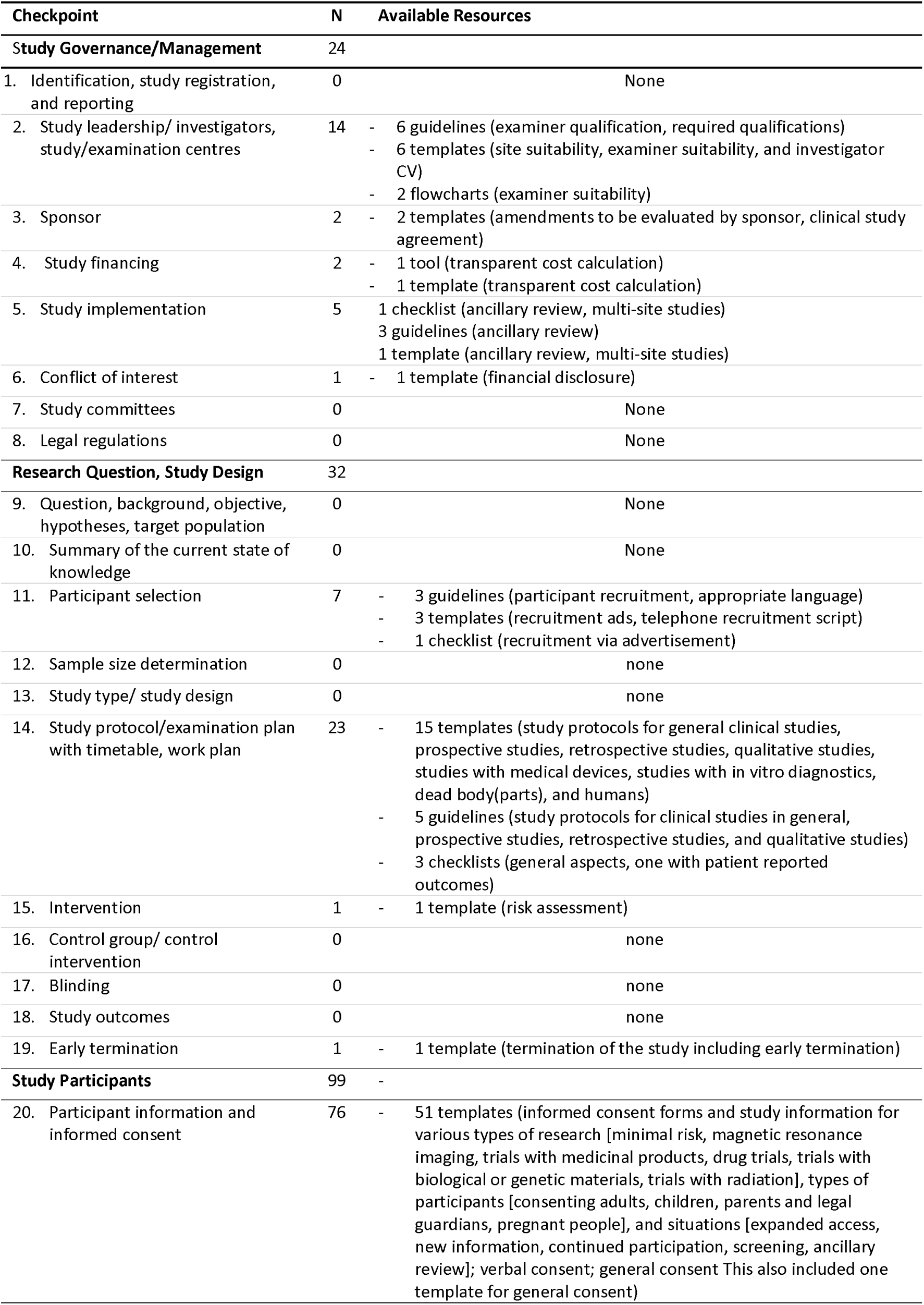
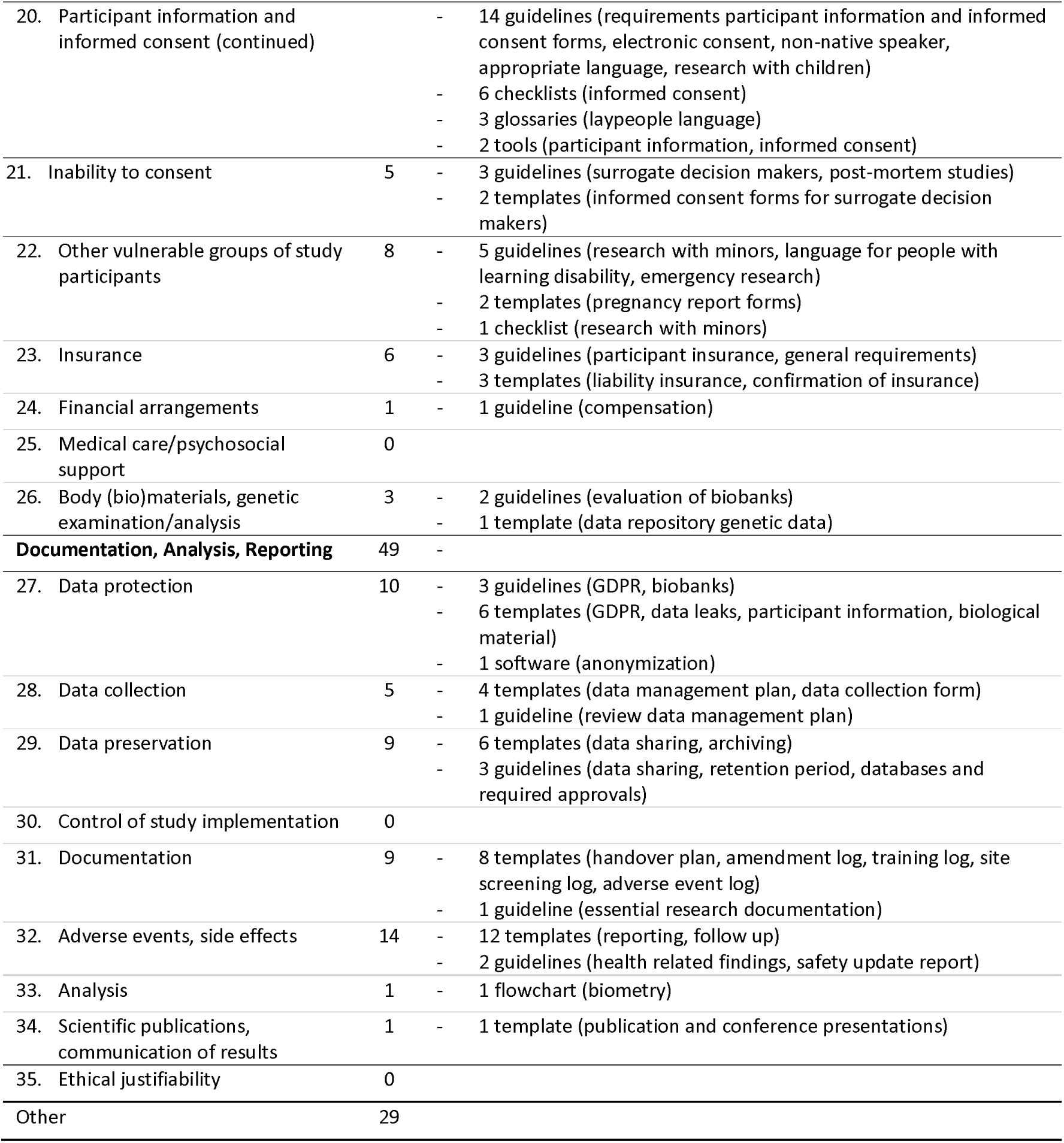
Extracted resources per checkpoint.

| Checkpoint | N | Available Resources |
| --- | --- | --- |
| **Study Governance/Management** | 24 |  |
| 1. Identification, study registration, and reporting | 0 | None |
| 2. Study leadership/ investigators, study/examination centres | 14 | - 6 guidelines (examiner qualification, required qualifications)<br>- 6 templates (site suitability, examiner suitability, and investigator CV)<br>- 2 flowcharts (examiner suitability) |
| 3. Sponsor | 2 | - 2 templates (amendments to be evaluated by sponsor, clinical study agreement) |
| 4. Study financing | 2 | - 1 tool (transparent cost calculation)<br>- 1 template (transparent cost calculation) |
| 5. Study implementation | 5 | 1 checklist (ancillary review, multi-site studies)<br>3 guidelines (ancillary review)<br>1 template (ancillary review, multi-site studies) |
| 6. Conflict of interest | 1 | - 1 template (financial disclosure) |
| 7. Study committees | 0 | None |
| 8. Legal regulations | 0 | None |
| **Research Question, Study Design** | 32 |  |
| 9. Question, background, objective, hypotheses, target population | 0 | None |
| 10. Summary of the current state of knowledge | 0 | None |
| 11. Participant selection | 7 | - 3 guidelines (participant recruitment, appropriate language)<br>- 3 templates (recruitment ads, telephone recruitment script)<br>- 1 checklist (recruitment via advertisement) |
| 12. Sample size determination | 0 | none |
| 13. Study type/ study design | 0 | none |
| 14. Study protocol/examination plan with timetable, work plan | 23 | - 15 templates (study protocols for general clinical studies, prospective studies, retrospective studies, qualitative studies, studies with medical devices, studies with in vitro diagnostics, dead body(parts), and humans)<br>- 5 guidelines (study protocols for clinical studies in general, prospective studies, retrospective studies, and qualitative studies)<br>- 3 checklists (general aspects, one with patient reported outcomes) |
| 15. Intervention | 1 | - 1 template (risk assessment) |
| 16. Control group/ control intervention | 0 | none |
| 17. Blinding | 0 | none |
| 18. Study outcomes | 0 | none |
| 19. Early termination | 1 | - 1 template (termination of the study including early termination) |
| **Study Participants** | 99 | - |
| 20. Participant information and informed consent | 76 | - 51 templates (informed consent forms and study information for various types of research [minimal risk, magnetic resonance imaging, trials with medicinal products, drug trials, trials with biological or genetic materials, trials with radiation], types of participants [consenting adults, children, parents and legal guardians, pregnant people], and situations [expanded access, new information, continued participation, screening, ancillary review]; verbal consent; general consent This also included one template for general consent) |
| 20. Participant information and informed consent (continued) |  | - 14 guidelines (requirements participant information and informed consent forms, electronic consent, non-native speaker, appropriate language, research with children)<br>- 6 checklists (informed consent)<br>- 3 glossaries (laypeople language)<br>- 2 tools (participant information, informed consent) |
| 21. Inability to consent | 5 | - 3 guidelines (surrogate decision makers, post-mortem studies)<br>- 2 templates (informed consent forms for surrogate decision makers) |
| 22. Other vulnerable groups of study participants | 8 | - 5 guidelines (research with minors, language for people with learning disability, emergency research)<br>- 2 templates (pregnancy report forms)<br>- 1 checklist (research with minors) |
| 23. Insurance | 6 | - 3 guidelines (participant insurance, general requirements)<br>- 3 templates (liability insurance, confirmation of insurance) |
| 24. Financial arrangements | 1 | - 1 guideline (compensation) |
| 25. Medical care/psychosocial support | 0 |  |
| 26. Body (bio)materials, genetic examination/analysis | 3 | - 2 guidelines (evaluation of biobanks)<br>- 1 template (data repository genetic data) |
| **Documentation, Analysis, Reporting** | **49** | - |
| 27. Data protection | 10 | - 3 guidelines (GDPR, biobanks)<br>- 6 templates (GDPR, data leaks, participant information, biological material)<br>- 1 software (anonymization) |
| 28. Data collection | 5 | - 4 templates (data management plan, data collection form)<br>- 1 guideline (review data management plan) |
| 29. Data preservation | 9 | - 6 templates (data sharing, archiving)<br>- 3 guidelines (data sharing, retention period, databases and required approvals) |
| 30. Control of study implementation | 0 |  |
| 31. Documentation | 9 | - 8 templates (handover plan, amendment log, training log, site screening log, adverse event log)<br>- 1 guideline (essential research documentation) |
| 32. Adverse events, side effects | 14 | - 12 templates (reporting, follow up)<br>- 2 guidelines (health related findings, safety update report) |
| 33. Analysis | 1 | - 1 flowchart (biometry) |
| 34. Scientific publications, communication of results | 1 | - 1 template (publication and conference presentations) |
| 35. Ethical justifiability | 0 |  |
| Other | 29 |  |

### Study Governance/Management

The topic “Study Governance/Management” covers the broad set of activities of principal investigators (PIs), sponsors or other staff involved in managing and governing a specific clinical study. Our search revealed supporting resources for most but not all the checkpoints grouped under this topic (5/8; 62.50%).

Support was offered regarding the qualifications of study management, investigators, and study sites (*n* = 14), the implementation of multi-site studies (n = 5) with a specific focus on ancillary review, the collaboration between study management and sponsors (n = 2)^5^, the transparent calculation and tracking of study financing (n = 2), and the reporting of conflict of interests (n = 1).

Our search revealed no resources to support the creation of an appropriate study title, study registration, and notification of authorities, the identification of relevant legal regulations, or the composition or creation of study committees.

### Research Question and Study Design

The topic “Research Question and Study Design” covers conceptual and methodological aspects of the study development and design. Our search revealed resources to support only the minority (4/11; 36.36%) of the checkpoints grouped under this topic.

Support was offered for the design and organization of study protocols (n = 23) including resources offering universal support or targeting specific study designs or data collection methods, the selection and specifically the recruitment of participants (n = 7), the risk assessment of an intervention in a clinical study involving therapies of medicinal products, and the reporting of trial termination to the responsible REC (n = 1).

Our search revealed no resources to support the other checkpoints relating to study design. Namely, the research question, background, objective, hypothesis, and target population, the summary of the current state of knowledge, sample size calculation, the study type and study design, the control group/intervention, blinding, and the study outcomes.

### Study Participants

The topic “Study Participants” covers aspects related to the safety and compensation of study participants. Our search revealed resources to support the majority (6/7; 85.71%) of the checkpoints grouped under this topic.

Specifically, support was offered for the preparation of participant information and informed consent forms (n = 74) including resources offering universal support or targeting specific study designs, participant types, or situations, the inclusion and treatment of members of vulnerable groups (n = 8), aspects related to insurance (n= 6), the development of informed consent strategies for participants that are incapable of providing consent (n = 5), the collection and storage of biological or genetic material (n = 3)^6^, and the financial compensation for participation (n = 1)^7^.

Our search did not yield results for resources to support setting up medical care or psychosocial support for study participants.

### Documentation, Analysis, Reporting

The topic “Documentation, Analysis, Reporting” covers aspects relating the documentation, storage, and sharing or data, methods, and results. Our search revealed resources to support the majority (7/8; 87.50%) of the checkpoints grouped under this checkpoint.

Specifically, support was offered for the documentation and reporting of adverse events and side effects (n = 14), the collection (n = 5) protection (n = 10), and preservation of data (n = 9), documentation purposes (n = 9), and the analysis (n = 1) and reporting (n = 1) of study results. Our search did not reveal any resources to control study implementation.

### Other

We extracted an additional 29 resources (n_English_ = 18, n_German_ = 11) that could not be grouped under one of the checkpoints. This included 12 checklists, nine guidelines, five templates, two tools, and one flowchart. The checklists concerned the required documents to be submitted for several types of studies, evaluation of the application or study protocol, continuing review, human research determination, project closure and the use of templates (from swissethics). Guidelines targeted the evaluation of applications, the obligation to report changes to the study protocol to the REC, human research determination, guidance how to apply through the online portal ethikPool or a compilation of international human research standards. The tools offered online training for clinical research ethics and the flowchart could be used to find answers to ethical questions that might arise during the research process.

## Discussion

### Resource Landscape

At the searched websites of umbrella organizations and RECs of major university hospitals we found a broad spectrum of resources supporting clinical researchers and members of RECs, most of which were templates, followed by guidelines and checklists.

Support varied between different checkpoints. Various types of resources are available for some aspects such as the preparation and assessment of participant information and consent form for different participant groups, subject areas (e.g., language, surrogate decision makers, etc.), and legal contexts, the preparation of study protocols, the assessment of qualifications of investigators and study sites, the reporting and documentation of adverse events, and data protection. Nonetheless, support is lacking in other areas (i.e., study design, analysis, and biometrics).

### Gaps

In some instances, there are reasonable explanations for a lack of resources. For example, umbrella organizations and RECs offered limited support for trial registrations, instead providing links to registry websites possibly under the assumption that registries should provide resources for trial registration, as the DRKS [Deutsches Register Klinischer Studien] and clinicaltrials.gov do (11,12)). However, this dispersion of information may pose a risk, making it time-consuming for applicants to find the necessary support and potentially explain some of the reported shortcomings of trial registrations (Thiele & Hirschfeld, 2022; Viergever).

Similarly, the searched websites did not provide resources to help researchers navigate the legal texts that govern many types of clinical trials. Instead, stakeholders frequently referred to the legal texts in full. As some legal requirements for clinical trials are reflected by checkpoints (e.g., qualification of the principal investigator), it is understandable that there were no resources specifically assigned to this checkpoint. Nonetheless, we believe that researchers could benefit from interpretive aids or checklists of the requirements arising from the legal regulatory framework.

This review also identified important gaps in the existing resources provided by the searched stakeholders in areas related to study design and biometrics. Few to no resources were found for these aspects. It is possible that this expertise resides outside of ethics. In contrast to other aspects, however, stakeholders did not refer to existing structures such as clinical study coordination systems or methodological and/or statistical consultation offered by university hospitals, which might be helpful for researcher to become aware of these services and seek them out. However, even then, the psychological barrier of a personal consultation may be high, which raises the question of the extent to which these services would be used. The need for simple, understandable, and easy-to-use resources on the topic of study design and biometrics for clinical research in humans, as for example available for animal research (e.g., the Experimental Design Assistant (15), remains.

Even for checkpoints with resources available, it remains open whether these indeed cover all relevant topic areas. For example, participant selection does not only concern the recruitment but also judgements regarding the representativeness of the participant pool, and appropriate in- and exclusion criteria. However, we did not find resources addressing these aspects of participant selection. Similarly, there is a scarcity of resources supporting the reporting of conflict of interest (CoI), despite tools developed by other stakeholders like journals and publishers to facilitate CoI statement creation (e.g., https://declarations.elsevier.com/).

### Opportunities

This project highlights opportunities for advancing and refining resources to support the clinical research ethics application process. The primary project output is a comprehensive overview of resources provided by key stakeholders. This overview can be used (1) for further tool and resource development and (2) a database for future research evaluating their quality and alignment with the needs of applicants and REC members.

Ultimately, these resources aim to enhance the quality of the ethical review process for all stakeholders, including applicants, REC members, and research participants. Presently, there are no established measures to evaluate quality and effectiveness of RECs (16). This is likely because the process of research ethics applications and approvals are based on ethical and regulatory standards that are subjectively applied (17). In the absence of effectiveness measures, tools, templates, checklists, and other resources can improve the clinical ethical review process. At the very least, systematic use of resources enhances efficiency of the review process and ensures compliance with regulatory and institutional policies, key quality indicators often mentioned by REC directors (Lynch et al., 2022). At best, the use of tools and resources for REC applications and decision-making procedures enables thoughtful engagement with the procedure of REC decision making and promotes consistent and robust standards (see for example Seykora et al., 2021).

### Limitations

Although this review has covered many prominent and relevant stakeholders, we recognize that our results do not cover all resources available to clinical researchers and REC members. Some information may be located on websites of stakeholders that were not included, other information may be shared internally, through password-protected pages, and other resources are likely available on websites of other organization types not included in this review. Additionally, the scope of our review was determined by thematic saturation. However, increased redundancy in some topics does not mean that there are not some resources on other topics on websites that we did not look at. We therefore highlight that this was a first scoping review exploring what kind of resources are provided at RECs and related umbrella organizations and should not be viewed as a comprehensive assessment of *all* available resources.

We caution against viewing our resource collection as a recommendation. In our qualitative analysis, we noticed markable variation in the depth and quality of different resources, but accuracy and timeliness were not examined. The qualitative value of the available resources remains unclear. Further detailed analysis and user testing are needed, but beyond the scope of this paper.

Lastly, we did not differentiate between supporting templates and forms required by the REC or (national) law. While we label forms as such in the amendment, we did not treat them as a separate category in the analysis, as forms can be considered supportive as they enhance transparency and the systematic nature of the research ethics application, facilitating understanding and compliance. This distinction may be explored in future research.

### Conclusion

This project provided an initial overview of the resources available to support applicants and REC members. We hope that this project will stimulate greater engagement with available resources and the identified thematic gaps, both in consultation with relevant stakeholder groups.

## Data Availability

All data produced are available online at https://osf.io/e7dmt/.

## Declarations

### Ethics approval and consent to participate

Not applicable

### Consent for publication

Not applicable

### Availability of data and materials

The datasets generated and analysed during the current study are available in the OSF repository, https://osf.io/e7dmt/, DOI 10.17605/OSF.IO/E7DMT

### Competing interests

The authors declare that they have no competing interests

### Funding

This work was conducted as part of the GUIDEME project funded by the German Federal Ministry of Education and Research (BMBF 01GP2208A). DS received the funding. The funder had no role in the study design, data collection and analysis, decision to publish, or preparation of the manuscript.

### Authors’ contributions

D.S.: Conceptualization, Funding acquisition, Methodology, Supervision, and Writing - review & editing.

M.-M.P.: Conceptualization, Data curation, Formal analysis, Investigation, Methodology, Project administration, Software, Visualization, and Writing - original draft.

## Acknowledgements

We thank Ulrike Grittner, Sebastian Graf von Kielmansegg, Lucas Augustin, Eva Winkler, and Annette Rid for their input and feedback at several stages of the project.

## List of Abbreviations

AKEK: Arbeitskreis Medizinischer Ethikkommissionen
IRB: Institutional Review Board
REC: Research Ethics Committee
WHO: World Health Organizationn

## Footnotes

1 The term IRB is more commonly used in America. In this article, we will use the term REC, which is more common in Europe.

2 Note that we have intentionally excluded textbooks and published papers from this list, as they are not as readily accessible and require a greater time commitment that is rarely achievable in application and review procedures for individual clinical studies.

3 Note that this limits the comparability of the number of resources between sources as we did not count the number of all resources available but the number of resources, not yet presented at previously extracted sources.

4 For seven stakeholders, we did not find any additional resources to extract.

5 Our search suggests that some RECs such as for example the University Hospital Bristol and University Hospital Birmingham offer institution specific resources regarding sponsorship. These did not meet our inclusion criteria but are presented in the data file on OSF under ‘other resources’.

6 Please note that the collection of biological materials was addressed in 13 additional resources including informed consent templates, insurance templates, and templates for participant information, which are grouped under different checkpoints.

7 However, financial arrangements and compensation was also mentioned in two additional resources. Guideline R134 grouped under participant recruiting explores the option of financial compensation and template R217 grouped under study financing has a section for financial arrangements.

## Notes

### Competing Interest Statement

The authors have declared no competing interest.

### Summary of Updates

This version of the manuscript has been revised following peer review comments. The updated version aims to convey the exploratory nature of this review more and highlight that the scope of the search is limited to a purposively selected sample of sources.

